# Local ancestry-aware genome-wide meta-analysis uncovers novel genetic loci for sickle cell disease nephropathy

**DOI:** 10.64898/2026.05.27.26354213

**Authors:** Melanie E. Garrett, Seyed Mehdi Nouraie, Roberto F. Machado, Victor R. Gordeuk, Mark T. Gladwin, NHLBI Trans-Omics for Precision Medicine Consortium, Marilyn J. Telen, Allison E. Ashley-Koch

## Abstract

In the United States, sickle cell disease (SCD) is a rare inherited hemoglobinopathy affecting about 100,000 individuals, mostly with African ancestry. SCD causes damage to multiple organ systems and SCD nephropathy (SCDN) is a common complication associated with early mortality. We previously performed a genome-wide association study (GWAS) for SCDN and identified a modest number of genome-wide significant loci. Here, we leveraged the ancestral composition of participants from two well-characterized adult SCD cohorts to boost statistical power and perform a local ancestry-aware GWAS for estimated glomerular filtration rate (eGFR), resulting in the identification of novel genome-wide significant loci within the African (AFR) and European (EUR) ancestral components of participants. Meta-analysis identified 12 significant genomic regions in the AFR tract, including *PPIL6*, *ARHGAP24*, *RAB11A*, and *STEAP3*, and 38 regions in the EUR tract, including *UBLCP1*, *ADAMTS6*, *JAZF1*, *MYO7B*, *MYO1C*, *PDGFA*, *GPC5*, *LRP1B*, *KANK1*, and *TRPV5*. The identified regions encompass genes affecting inflammation, extracellular matrix (ECM) integrity, iron metabolism, magnesium ion homeostasis, B cell apoptosis, tumor necrosis factor (TNF) production, and estrogen signaling. Many of these genes and pathways are important not only for renal function, but also for SCD biology, providing additional support for the hypothesis that SCDN pathophysiology is unique from other forms of kidney disease. This study represents the largest local ancestry-aware analysis of SCDN to date, furthers our understanding of the genetic risk factors underlying SCDN, and proposes new targets that could be useful for the early identification and treatment of kidney dysfunction in SCD patients.

**KEY POINTS:**

- Novel application of local ancestry-aware GWAS in two SCD cohorts identified distinct eGFR loci from the African and European ancestries
- This application increased power to detect new loci with biologic functions important to renal function, as well as sickle cell disease

## INTRODUCTION

In the United States (US), sickle cell disease (SCD) is a rare inherited hemoglobinopathy affecting about 100,000 individuals of primarily African ancestry^1^. A single base pair mutation in the beta globin gene (*HBB*) produces sickle hemoglobin (HbS), which polymerizes under hypoxic conditions, causing sickling of red blood cells, vaso-occlusion and subsequent ischemia, hemolytic anemia, organ damage, and reduced survival^2–4^. The renal system is commonly affected by SCD, and, importantly, we have shown that renal dysfunction is associated with early mortality^3^. SCD nephropathy (SCDN) occurs in 5-30% of patients as a result of intravascular hemolysis, vaso-occlusion and decreased blood flow to the kidneys, leading to renal hypertrophy, hyposthenuria, proximal tubular dysfunction, glomerular abnormalities, and ultimately renal failure^3,5–8^. Nearly 70% of patients develop micro- or macroalbuminuria and 4-12% of adults with SCD progress to end-stage renal disease (ESRD)^6,9,10^. Early detection of SCDN is crucial to identify at-risk patients prior to significant kidney damage but remains challenging, as current practice relies on relatively late-stage disease markers, such as proteinuria and decreased glomerular filtration rate (GFR). Identification of genetic modifiers of SCDN could allow clinical risk-stratification of patients before significant kidney dysfunction occurs and forestall further progression to ESRD and premature death.

Large genome-wide association studies (GWAS) of chronic kidney disease (CKD) and other renal phenotypes in non-SCD cohorts have identified many disease-associated loci^11–14^. However, polygenic risk scores (PRS) generated from the largest of these^11,12^ are poor predictors of SCDN (R^2^<0.01)^15^, likely due to the unique biological mechanisms underlying SCDN and the overwhelmingly European ancestry (>97%) from which the PRSs were generated. We previously performed discovery array-based GWAS for SCDN in two SCD cohorts (Outcome Modifying Genes in Sickle Cell Disease [OMG-SCD] and Walk-Treatment of Pulmonary Hypertension and Sickle Cell Disease with Sildenafil Therapy [Walk-PHaSST]) and identified three genome-wide significant (GWS) loci for estimated GFR (eGFR) that are independent of the well-known *APOL1* locus^15^. These loci are distinct from those reported in the non-SCD European studies and have biological relevance to both renal function and SCD^15^, highlighting the need for continued nephropathy research in SCD cohorts of African ancestry. Since that work, several GWAS of kidney phenotypes have been performed in predominantly African and Hispanic/Latino ancestry cohorts^16–18^; these identified the SCD causal variant, *HBB* rs334, as an independent risk factor, likely driven by the inclusion of participants with sickle trait (heterozygous for rs334) and underscoring the SCD variant as a driver of kidney dysfunction.

Genetic modifier studies of SCD in the US are notoriously underpowered due to the rarity of the disease compared to other regions of the world^19^. Thus, ascertaining the number of patients necessary to perform well-powered studies is challenging. OMG-SCD and Walk-PHaSST are two of the largest adult SCD cohorts in the US for which whole genome sequencing (WGS) data are available as a part of the National Heart, Lung, and Blood Institute (NHLBI) Trans-Omics for Precision Medicine (TOPMed) program. Here, we endeavored to uncover novel genetic loci for SCDN by applying statistical approaches that increase statistical power by harnessing the ancestral composition (African and European) of the SCD patients. Specifically, we applied admixture mapping methods leveraging the idea that disease-associated variants may display disparate allele frequencies and effects across ancestral populations^20^. Recently, an improved method that generates ancestry-specific summary statistics regardless of whether ancestral allele frequencies differ by disease, called *Tractor*, was developed^21^. This method uses local ancestry at each locus genome-wide to uncover genetic signals that are present in only one ancestral population and are therefore unidentifiable in standard GWAS. Utilizing *Tractor*, we performed a local ancestry-aware GWAS for estimated GFR (eGFR) in OMG-SCD and Walk-PHaSST and did indeed identify novel SCDN loci previously undetectable with standard approaches.

## MATERIALS AND METHODS

### Participants and Endpoints

Adult (age ≥18 years) SCD participants with genetic data and clinical variables necessary for calculating eGFR were selected from two previously described cohorts for inclusion in this study. 598 participants were included from OMG-SCD^22^, a multi-center cohort from the southeastern US, ascertained for a cross-sectional observational study to investigate genetic modifiers of SCD. 368 participants were included from the Walk-PHaSST study^23^, a clinical trial of sildenafil, for which participants were enrolled during the screening phase at centers in the US and United Kingdom. All participants gave informed consent and study protocols were approved by local institutional review boards or ethics committees.

Participants provided blood samples for genetic analyses at study enrollment, and a variety of clinical variables were obtained at steady state. Baseline eGFR was calculated using the CKD-EPI-2021 equation, based on serum creatinine, age, and sex, with no race correction^24^. To control for the effect of hemoglobin beta genotype, participants were grouped into more severe (SS and Sβ0 thalassemia) and less severe (SC and Sβ+ thalassemia) disease.

### Whole genome sequencing

DNA extracted from whole blood underwent ∼30X WGS as part of the NHLBI TOPMed program at Baylor College of Medicine. All samples in this study were included in TOPMed freeze 8, and data processing and primary quality control (QC) were performed at the Informatics Research Center (IRC) at the University of Michigan, details of which have been previously described^25^. Briefly, raw sequencing reads were aligned to the GRCh38 reference assembly, joint variant calling was performed including approximately 158,000 freeze 8 samples, and identified variants were filtered using a support vector machine (SVM). In total, 653 unique OMG-SCD and 437 Walk-PHaSST samples were sequenced as part of the TOPMed program (**Figure S1**). Samples were inspected for sex mismatches, pedigree errors, and concordance with previous genotyping array data. In this analysis, only variants with a minimum read depth of 10X were considered. Additional QC steps were imposed on OMG-SCD and Walk-PHaSST separately, including removal of variants missing for >15% of samples or with minor allele frequency (MAF) <1% and restriction to biallelic single nucleotide polymorphisms (SNPs). Finally, identity-by-descent (IBD) estimates were generated in PLINK^26^ after extracting linkage disequilibrium (LD)-pruned SNPs with MAF >20%. One sample from pairs with IBD estimates >0.25 (second degree relatives) was randomly excluded in Walk-PHaSST (N=31); no second degree or higher relationships were identified in OMG-SCD.

### Statistical analysis

Differences in participant characteristics between OMG-SCD and Walk-PHaSST were analyzed in R using chi-square tests (*chisq.test*) for categorical variables and generalized linear models (*glm*) for continuous variables. In each cohort, principal component (PC) analysis was performed with EIGENSOFT v10^27^ using LD-pruned SNPs with MAF >20%. Eight samples for OMG-SCD and ten samples for Walk-PHaSST were classified as PC outliers and removed. Two PCs were sufficient to account for population substructure in OMG-SCD; five PCs were necessary in Walk-PHaSST according to visual inspection of scree plots (**Figure S2**). Estimates of global European admixture were calculated for each participant with the linkage model in STRUCTURE^28^, utilizing over 4000 ancestry informative markers^29^. Two Walk-PHaSST participants had global European admixture percentages >80% and were excluded.

Local ancestry-aware GWAS for eGFR was conducted in each cohort separately, controlling for sex, age, SCD severity, and global European admixture percentage. Local-ancestry inference (LAI) was performed using RFMix^30^ to obtain ancestry estimates at each locus genome-wide. To confirm LAI estimates were unbiased, we compared average local ancestry to global estimates from STRUCTURE and observed very high correlation in OMG-SCD and Walk-PHaSST (r=0.979 and 0.990, respectively; **Figure S3**). Painted karyogram plots were generated using publicly available code (https://github.com/armartin/ancestry_pipeline); plots for selected participants with varying global European admixture percentages are shown in **Figure S4**. LAI estimates were then used to perform local ancestry-aware GWAS using *Tractor*^21^, which generates results for the African (AFR) and European (EUR) ancestry tracts separately. For comparison, a standard eGFR GWAS was performed in each cohort using PLINK and controlling for sex, age, SCD severity, and genotype PCs, as described above.

Fixed effects meta-analysis was accomplished with METAL^31^, combining results across cohorts for the African ancestry-specific GWAS (N=8,900,470 SNPs), the European ancestry-specific GWAS (N=4,669,672 SNPs), and the standard GWAS (N=10,916,045 SNPs). Using the standard genome-wide threshold, SNPs with p-value<5×10^-8^ were considered significant. Random effects meta-analysis using R *metafor*^32^ was performed for SNPs significant in the fixed effects model but showing heterogeneity across cohorts (Cochran’s Q test p<0.05); these SNPs were excluded if the random effects meta-analysis was not significant (p<0.05). Quantile-quantile (QQ) and Manhattan plots were produced using R qqman^33^, and variant annotations were queried using ANNOVAR^34^.

Following liftOver^35^ to genomic build 37, we utilized FUMA^36^ to perform post-GWAS annotation of GWS regions, including metrics of predicted deleteriousness (CADD scores)^37^ and identification of genes mapped to the GWS regions. To investigate additional evidence for regulatory function, candidate SNPs were queried in NephQTLv2^38^, a public database of expression quantitative trait loci (eQTL) in glomerular and tubulointerstitial human kidney tissue from subjects with nephrotic syndrome, and were assessed for impact on transcription factor (TF) binding using QBiC-Pred^39^.

Finally, gene-based analysis was performed using MAGMA^40^ as implemented in FUMA, and false discovery rate (FDR) q-values were calculated using the R package *qvalue*^41^. Gene set enrichment analysis was performed using ShinyGo v.0.85.1^42^. Two gene sets were tested for enrichment: genes with p<0.01 from the AFR tract and EUR tract MAGMA gene-based analyses. All human genes were considered for the background gene set. Specifically, we tested for enrichment of KEGG (Kyoto Encyclopedia of Genes and Genomes) pathways and the three main GO (Gene Ontology) term categories: biological processes, cellular components, and molecular function. Terms with FDR q<0.05 were deemed significant, and lollipop plots ordered by fold enrichment were generated in GraphPad Prism v11 (GraphPad Software, Boston, MA).

## RESULTS

### Study participants

Participant characteristics are shown in **Table 1**. OMG-SCD participants were younger (33.41 vs. 38.34 years old, p=3.42×10^-9^), had a higher proportion of hemoglobin SS or Sβ^0^ disease (80.77% vs. 74.18%, p=6.43×10^-3^), and had a lower average proportion of European admixture (0.156 vs. 0.178, p=3.83×10^-3^) than Walk-PHaSST participants. OMG-SCD participants also displayed higher mean eGFR compared to Walk-PHaSST participants (111.44 mL/min/1.73m^2^ vs. 104.22 mL/min/1.73m^2^, p=7.20×10^-4^); however, once we controlled for age, there was no mean difference in eGFR (p=0.619).

**Table 1.** Participant characteristics by cohort.

|  | OMG-SCD<br>(N=598) | Walk-PHaSST<br>(N=368) | p-value |
| --- | --- | --- | --- |
| Mean age, years (SD) | 33.41 (12.30) | 38.34 (12.72) | 3.42x10<sup>-9</sup> |
| % Female | 54.85% | 54.89% | 0.99 |
| % HbSS/Sβ<sup>0</sup> | 80.77% | 74.18% | 6.43x10<sup>-3</sup> |
| Mean global EUR admixture,<br>proportion (SD) | 0.156 (0.091) | 0.178 (0.138) | 3.83x10<sup>-3</sup> |
| Mean eGFR, mL/min/1.73m<sup>2</sup> (SD) | 111.44 (32.42) | 104.22 (31.63) | 7.20x10<sup>-4</sup> |
| Mean age-adjusted eGFR,<br>mL/min/1.73m<sup>2</sup> (SD) | 108.37 (1.02) | 109.20 (1.31) | 0.619 |

SD=standard deviation, EUR=European, eGFR=estimated glomerular filtration rate

### Local ancestry-aware GWAS

Local ancestry-aware GWAS for eGFR was performed and genomic inflation was well controlled (**Figure S5**). We identified 12 significant genomic regions in the AFR tract encompassing 109 SNPs and 18 mapped genes **(Figure 1A**; **Table 2**). The most significant AFR tract SNP was rs35371039 (p=3.72×10^-14^), an intergenic SNP on chromosome 4. For the EUR tract, we identified 38 significant regions encompassing 269 SNPs and 53 mapped genes (**Figure 1B**; **Table 3**). The most significant EUR tract SNP was rs140629823 (p=3.64×10^-13^), an intergenic SNP on chromosome 14. Results for OMG and Walk-PHaSST prior to meta-analysis are shown in **Table S1-2** and include ancestry-specific MAF, local ancestry proportion, and eGFR effect sizes and p-values. None of the eGFR-associated regions in the AFR tract were the same as those in the EUR tract, suggesting that there are highly differing allele frequencies and/or large effect sizes differences in the AFR and EUR ancestries of these participants.

**Figure 1.**
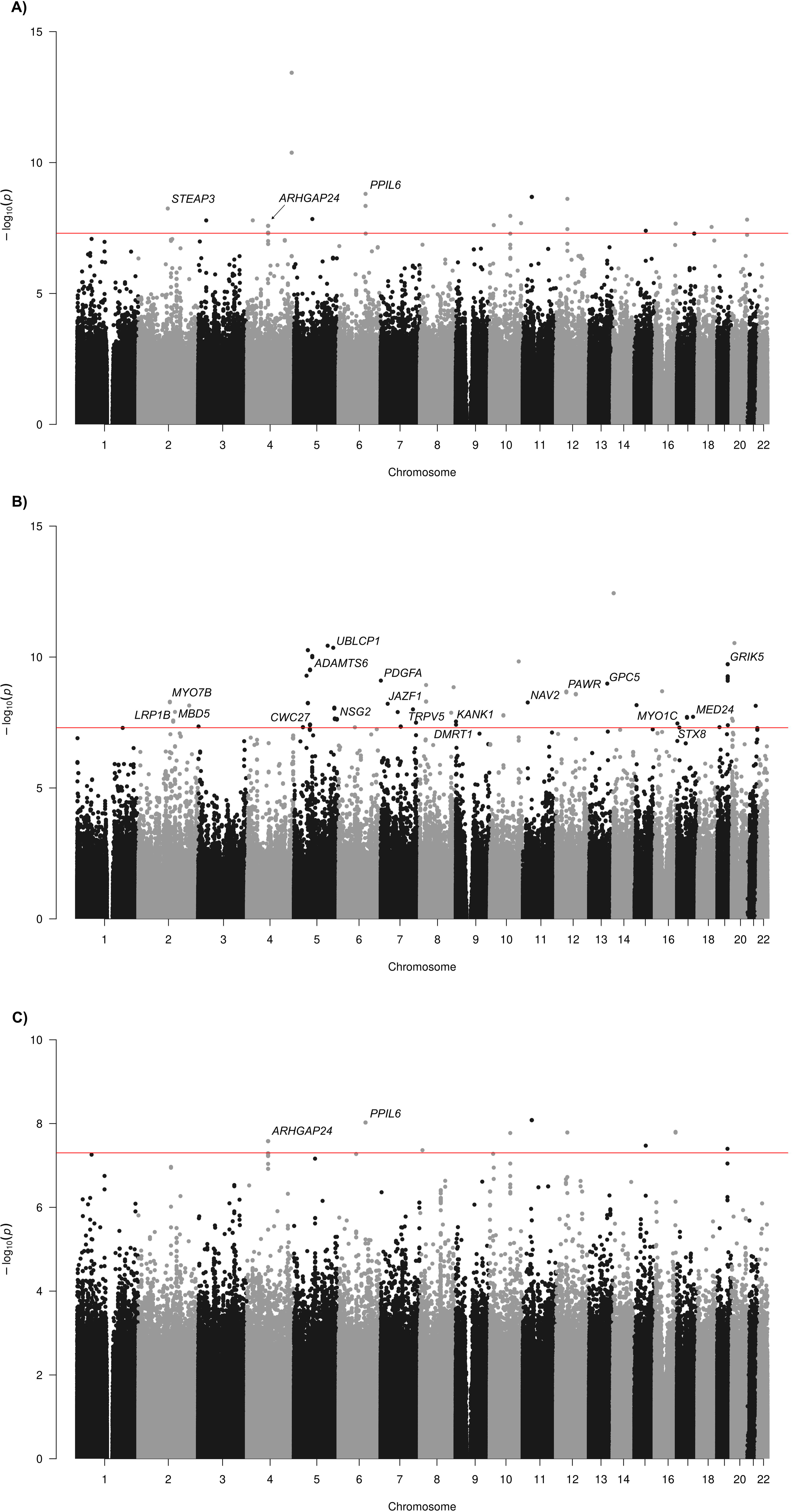
Manhattan plots for eGFR using local ancestry-aware GWAS results for the A) AFR and B) EUR tracts compared to the C) standard GWAS model. The genome-wide significance threshold (p=5×10^-8^) is depicted by the red line. SNPs residing in genes are labeled with the gene symbol.

**Table 2.**
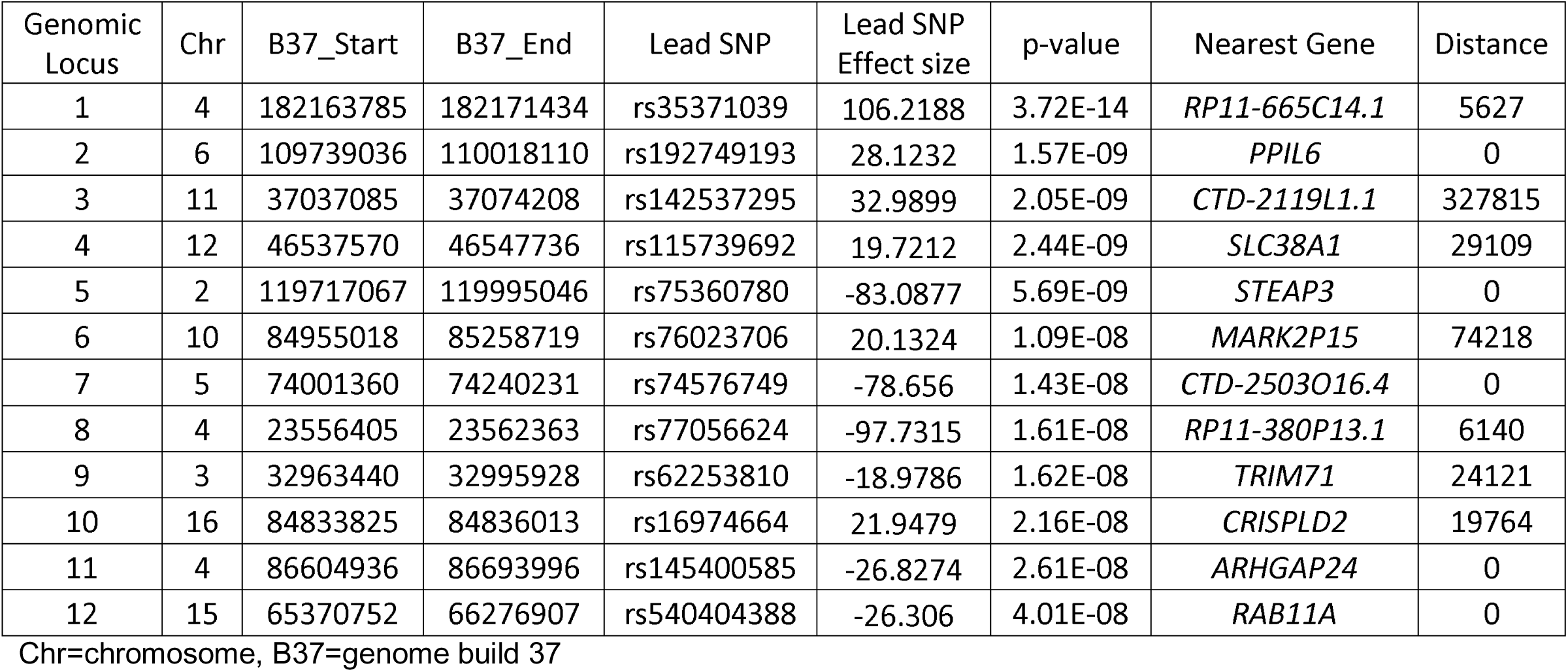
African tract genome-wide significant regions.

| Genomic Locus | Chr | B37_Start | B37_End | Lead SNP | Lead SNP Effect size | p-value | Nearest Gene | Distance |
| --- | --- | --- | --- | --- | --- | --- | --- | --- |
| 1 | 4 | 182163785 | 182171434 | rs35371039 | 106.2188 | 3.72E-14 | *RP11-665C14.1* | 5627 |
| 2 | 6 | 109739036 | 110018110 | rs192749193 | 28.1232 | 1.57E-09 | *PPIL6* | 0 |
| 3 | 11 | 37037085 | 37074208 | rs142537295 | 32.9899 | 2.05E-09 | *CTD-2119L1.1* | 327815 |
| 4 | 12 | 46537570 | 46547736 | rs115739692 | 19.7212 | 2.44E-09 | *SLC38A1* | 29109 |
| 5 | 2 | 119717067 | 119995046 | rs75360780 | -83.0877 | 5.69E-09 | *STEAP3* | 0 |
| 6 | 10 | 84955018 | 85258719 | rs76023706 | 20.1324 | 1.09E-08 | *MARK2P15* | 74218 |
| 7 | 5 | 74001360 | 74240231 | rs74576749 | -78.656 | 1.43E-08 | *CTD-2503O16.4* | 0 |
| 8 | 4 | 23556405 | 23562363 | rs77056624 | -97.7315 | 1.61E-08 | *RP11-380P13.1* | 6140 |
| 9 | 3 | 32963440 | 32995928 | rs62253810 | -18.9786 | 1.62E-08 | *TRIM71* | 24121 |
| 10 | 16 | 84833825 | 84836013 | rs16974664 | 21.9479 | 2.16E-08 | *CRISPLD2* | 19764 |
| 11 | 4 | 86604936 | 86693996 | rs145400585 | -26.8274 | 2.61E-08 | *ARHGAP24* | 0 |
| 12 | 15 | 65370752 | 66276907 | rs540404388 | -26.306 | 4.01E-08 | *RAB11A* | 0 |

Chr=chromosome, B37=genome build 37

**Table 3.**
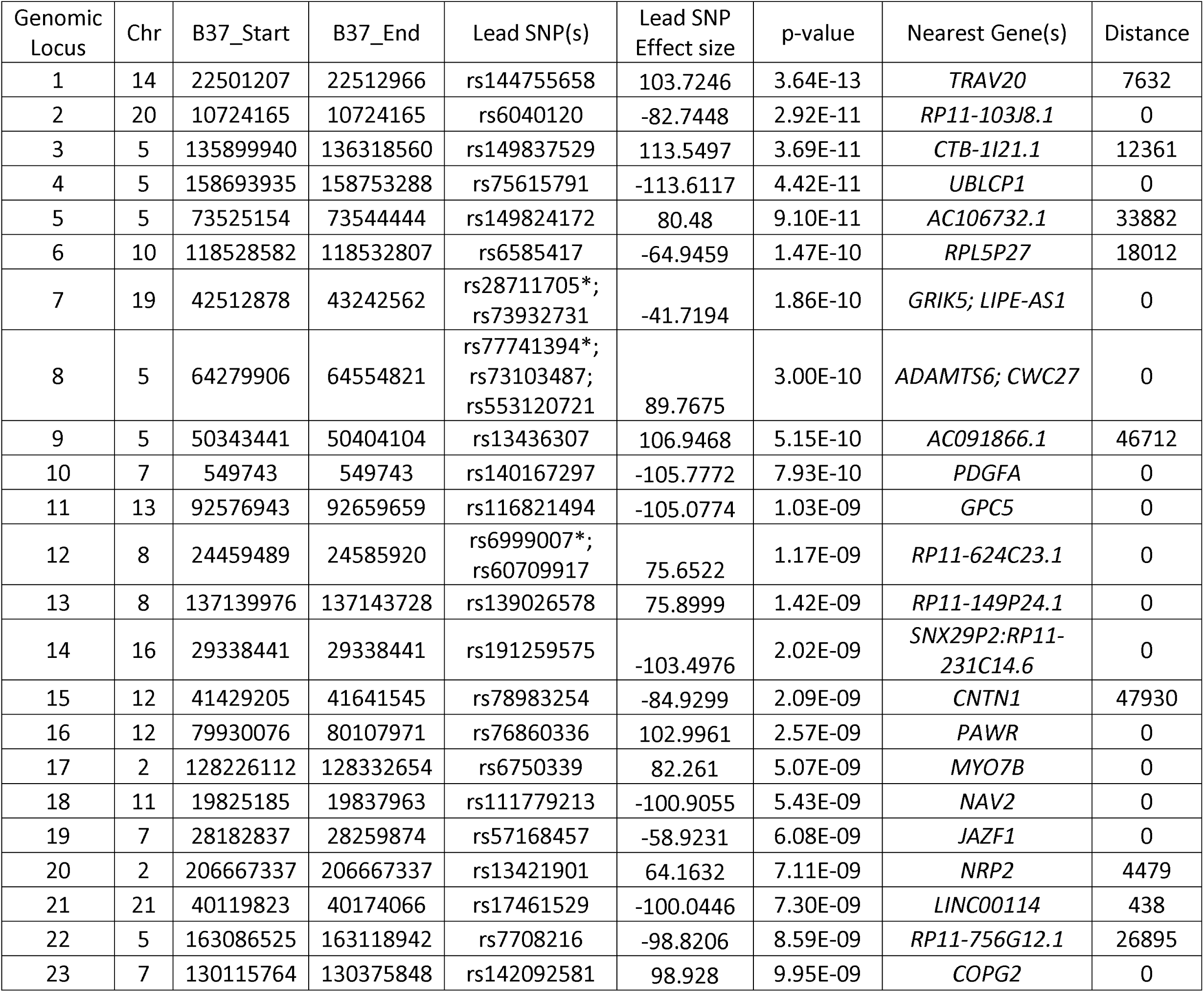
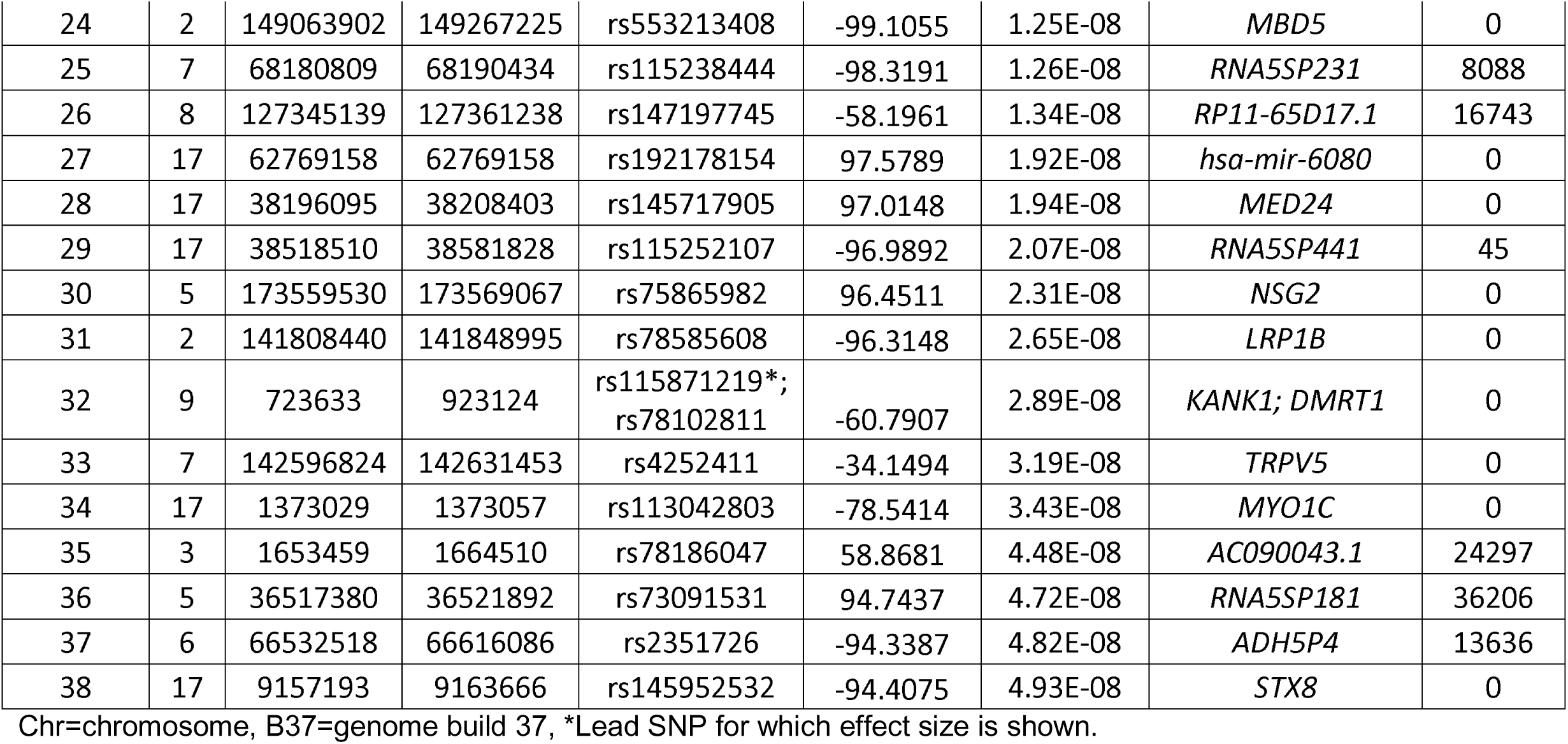
European tract genome-wide significant regions.

Many SNPs in the eGFR-associated regions displayed CADD scores above 10, indicating they rank among the top 10% most deleterious variants genome-wide (**Table S3-4**). For the AFR tract, the highest CADD score (15.76) belonged to rs71619055, located in the most significant eGFR-associated region. For the EUR tract, rs78816499, a synonymous variant in *POU2F2,* had the highest CADD score (17.05). Many of the genes mapped to GWS regions are likely loss-of-function intolerant (pLI>0.99), including *FAM169A* for the AFR tract GWAS and *LRP1B, CNTN1, CIC, MBD5, ADAMTS6, MEST, NAV2, GSK3A, TOP2A, and ETS2* for the EUR tract GWAS (**Table S5-6**).

We also observed evidence that SNPs in eGFR-associated regions may exhibit regulatory function. Utilizing kidney-specific eQTL data from NephQTL, we note that rs75360780, the lead SNP in AFR tract genomic locus 5, is an eQTL for *STEAP3*, in tubulointerstitium (p=6.66×10^-5^, q=0.015). EUR tract genomic locus 6 harbors eQTLs for *HSPA12A* in glomeruli including lead SNP rs6585417 (p=1.81×10^-5^, q=0.0109), rs6585416 (p=4.31×10^-5^, q=0.0225), and rs7093563 (p=1.24×10^-4^, q=0.0522). Aside from changes in gene expression, many SNPs in eGFR-associated regions either disrupt or activate TF binding (**Table S3-4**), including NFκB and other associated factors (REL, RELA, RELB), kruppel-like factors (KLFs) including KLF4, forkhead box members FOXK1, FOXO3, and FOXP2, NFATC1, POU4F1, TFEB, and UNCX, which have been previously associated with CKD and suggested therapeutic targets.

### Standard GWAS

For comparison, we performed a standard eGFR GWAS and identified seven significant regions comprising 119 SNPs and seven mapped genes (**Figure 1C; Table S7-9**). Of the 12 regions identified in the AFR tract, six were also identified in the standard GWAS. Of the 109 GWS and LD SNPs in the AFR tract, 107 were at least nominally significant in the standard GWAS (98.2% overlap). By contrast, none of the 38 significant EUR tract regions were significant in standard GWAS. Of the 269 GWS and LD SNPs in the EUR tract, only 72 were at least nominally significant in the standard GWAS (26.8% overlap). These observations are consistent with the degree to which these two ancestries are present in our cohorts.

### Gene-based analysis

Gene-based analysis identified 29 genes associated with eGFR in the EUR tract (**Figure 2**; **Table S10**; FDR q<0.05). The most significant gene was *LINC01948*, a long non-coding RNA on chromosome 5 (p=1.43×10^-9^), followed by *ACOXL* and *RARA-AS1* (p=1.94×10^-7^ and p=2.78×10^-7^, respectively). None of the genes identified in the AFR tract surpassed correction for multiple testing.

**Figure 2.**
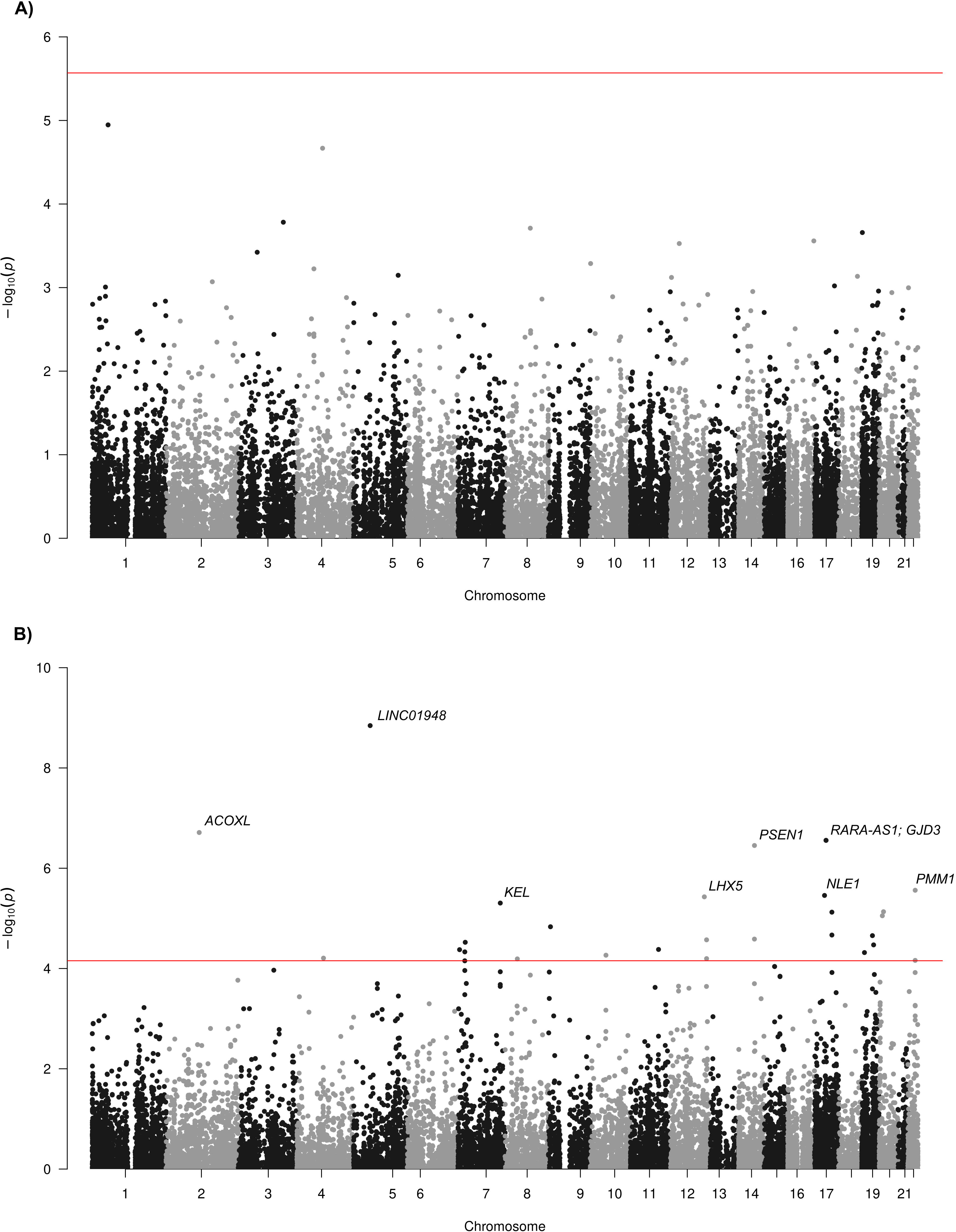
Manhattan plots for gene-based eGFR association results. AFR tract results are shown in panel A) and EUR tract results are shown in panel B). The horizontal red line demarcates FDR q-value=0.05. Genes with FDR q-value≤0.01 are labeled with the gene symbol.

Gene set enrichment of the top EUR tract genes (p<0.01) identified seventeen significant GO biological process terms, including intracellular magnesium ion homeostasis, negative regulation of B cell apoptosis, sulfur amino acid metabolism, and tumor necrosis factor (TNF) regulation (37.5, 17.9, 10.7 and 3.8-fold enrichment, respectively; **Figure 3**, **Table S11**). We also observed a 4.5-fold enrichment of genes in the estrogen signaling pathway when querying KEGG pathways. There was no enrichment of GO terms or KEGG pathways for the top AFR tract genes.

**Figure 3.**
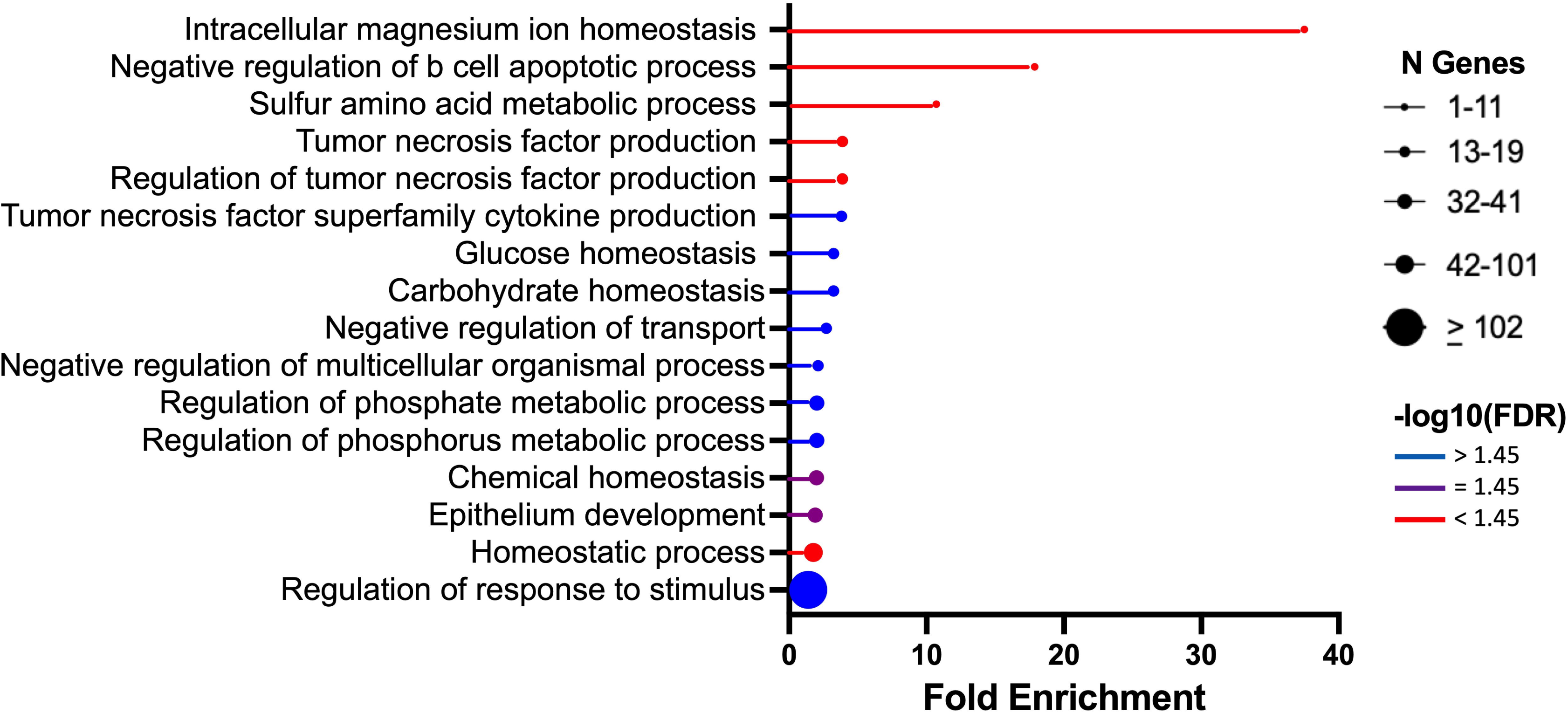
Lollipop plot of significant (FDR q<0.05) gene ontology biological process terms from gene set enrichment analysis of EUR tract gene-based results

## DISCUSSION

We have performed the first local ancestry-aware GWAS for SCDN, thereby identifying genome-wide significant variants and genes independent from one another in the African and European ancestral components. This analysis, by uncovering loci previously undetected by standard GWAS approaches, demonstrates the power of applying ancestry-aware techniques to admixed populations.

Standard GWAS, performed for comparison, showed the expected strong overlap with the AFR tract GWAS, since the SCD participants in this study are 83% African on average. Therefore, most signals identified in the standard GWAS are attributable to the AFR genetic component of each participant. In fact, only two variants identified in the AFR tract were not at least nominally significant in the standard GWAS (107/109 variants).

Genes with signals in both the AFR tract and the standard GWAS included *PPIL6*, *ARHGAP24*, and *RAB11A*. Peptidylprolyl Isomerase Like 6 (*PPIL6*) belongs to the cyclophilin family of peptidyl-prolyl cis/trans isomerases (PPIase), but its function is not well characterized^43^. However other related PPIases, cyclophilin A and Pin1, are involved in the onset and progression of kidney diseases via regulation of fibrosis, oxidative stress, and inflammation^44,45^. Both are proposed biomarkers of kidney disease and potential therapeutic targets. Another SNP in the *PPIL6* GWS region, rs187137934 in nearby *AK9*, disrupts binding of several KLF TFs, which are crucial for kidney function. The rs187137934 T allele, associated with low eGFR in our cohort, prevents KLF4 binding, a protective factor that modulates renal fibrosis by regulating inflammation^46,47^. We also identified GWS variants in *ARHGAP24*, a Rho GTPase that maintains podocyte structure and glomerular basement membrane integrity. Mutations in *ARHGAP24* have been identified in patients with steroid-resistant nephrotic syndrome (SRNS)^48^ and focal segmental glomerulosclerosis (FSGS)^49^. Moreover, five of the *ARHGAP24* SNPs we identified have CADD scores above 10, indicating a high likelihood of deleteriousness. Finally, we identified GWS variants in *RAB11A*, which encodes a small GTPase necessary for vesicle trafficking in the renal proximal tubules. *RAB11A* is loss-of-function intolerant (pLI=0.98), and its dysfunction has been implicated in animal models of renal ischemia-reperfusion injury, a major cause of acute kidney injury (AKI) and risk factor for CKD^50,51^.

Despite overwhelming concordance between the AFR tract and standard GWAS, we were able to identify novel associations in *STEAP3* by leveraging local ancestry. *STEAP3* encodes a metalloreductase involved in iron metabolism and affects ferroptosis, a form of iron-dependent cell death. STEAP3 is upregulated in renal cell carcinoma (RCC)^52,53^ and may play a role in kidney disease through regulation of iron homeostasis^54,55^. The *STEAP3* variant we identified is an eQTL in kidney tubulointerstitium of patients with nephrotic syndrome, suggesting that rs75360780 acts through regulation of STEAP3 expression. Notably, this region was not associated in the standard GWAS, highlighting the boost in statistical power obtainable by performing local ancestry-aware analysis.

While most eGFR-associated regions in the AFR tract and the standard GWAS were the same, the regions identified in the EUR component were distinct. The most significantly associated variant within a gene was rs75615791 in *UBLCP1*, a ubiquitin phosphatase that dephosphorylates the 26S proteasome, thereby dysregulating protein degradation. When impaired in the podocytes, this process can lead to CKD^56,57^. Moreover, the A allele of rs75615791, associated with low eGFR in our cohorts, prevents binding of KLF4, a renoprotective TF^46,47^. We identified SNPs in a disintegrin and metalloprotease with thrombospondin motifs (ADAMTS) 6 (*ADAMTS6*), whose expression is upregulated in proximal renal tubular epithelial cells in Fabry disease, which is characterized by CKD^58^. A prediction model comprised of ADAMTS proteins including ADAMTS6 strongly predicts survival in clear cell RCC^59^. Of note, we previously reported association of *ADAMTS7* SNPs with proteinuria in this cohort^15^. SNPs in *JAZF1* were associated with eGFR in the EUR tract; previous reports have identified changes in *JAZF1* DNA methylation at cg23597162 associated with CKD and with differences pre- and post-kidney transplant^60,61^. Interestingly, Schlosser and colleagues^60^ note that the cg23597162 association showed significant heterogeneity between European and African ancestry cohorts, consistent with our finding in the EUR tract only. Two different myosin genes are implicated in the EUR tract: *MYO7B* encodes a protein expressed in brush border microvilli of kidney epithelial cells, disruption of which results in reduced GFR, and *MYO1C*, variants in which have been associated with proteinuric kidney disease^62–64^. Many other genes identified here have been previously associated with various types of kidney disease or dysfunction, including *PDGFA*, *GPC5*, *LRP1B*, *KANK1*, and *TRPV5*^15,65–70^. Taken together, we observed enrichment of eGFR-associated EUR tract genes involved in intracellular magnesium ion homeostasis^71^, regulation of B cell apoptosis^72^, TNF production^73^, and estrogen signaling^74^, all of which have been previously implicated in renal dysfunction.

Many genes and pathways identified here are important not only for proper kidney function but also for SCD pathophysiology, either through involvement in pathologic mechanisms or associations with SCD-related complications. Polymerization of mutated hemoglobin S (HbS) due to oxygen deprivation results in red blood cell sickling, hemolytic anemia, and oxidative stress that together produce a chronic inflammatory state. Cells containing primarily HbS undergo cytoskeleton changes leading to structural instability, abnormal ion fluxes due to altered membrane permeability, and increased reactive oxygen species (ROS) production^75,76^. Therefore, the associations we observed in *ARHGAP24*, which encodes a Rac1-specific Rho GTPase that regulates inflammation through ROS production, may be particularly relevant for kidney dysfunction in the context of SCD^77^. The associations in *ADAMTS6*, a metalloprotease involved in extracellular matrix (ECM) remodeling through fibrillin protein cleavage and whose expression is associated with increased transforming growth factor-beta (TGF-β) activation^78^ are interesting because HbS red cells display increased adhesion to ECM proteins. Moreover, plasma levels of other matrix metalloproteases, like MMP9, as well as TGF-β, are increased in SCD patients and are associated with vaso-occlusive crises and inflammation^79,80^. Repeated hemolysis and anemia, as well as recurrent blood transfusions to replace HbS red cells and treat SCD complications, can contribute to iron overload, another consequence of SCD. Therefore, the association we saw in *STEAP3*, a metalloreductase involved in iron metabolism and ferroptosis, is particularly intriguing. Indeed, Steap3 is upregulated in the liver of hemolysis-induced Townes SCD mice compared to littermate controls and may be a useful biomarker of chronic organ injury in SCD^81^. Finally, we identified eGFR-associated variants in *JAZF1*, which has been associated with hemoglobin F (HbF) levels in patients with SCD and β-thalessemia^82,83^. These data show many of the eGFR-associated regions we identified harbor genes and pathways integrally related to biological processes underlying SCD.

As previously described^21^, power gains from *Tractor* are biggest when large effect size differences exist and/or the minor allele frequencies differ greatly between ancestries. The most extreme cases occur when the risk allele in one ancestry is monomorphic in the other ancestry or when the effect is restricted to the haplotype of the less frequent ancestral population. The large majority of AFR tract GWS SNPs we identified were monomorphic in the EUR tract (84.4% in OMG-SCD and 78.9% in Walk-PHaSST; **Table S1**), explaining why the AFR tract results were largely stronger than those from the standard GWAS. Conversely, the EUR tract associations appear to be the result of very different effect sizes in the AFR and EUR components (**Table S2**). For these loci, the association with eGFR only exists on the EUR haplotype, which accounts for approximately 15% of the global ancestral composition of the patients in our cohorts, and elucidates why these loci have been previously undetectable in standard GWAS.

We previously reported three SNPs associated with eGFR in the same SCD cohorts using imputed array-based GWAS^15^. In the current analysis, both rs1968911 in *LRP1B* and rs4903539 in *linc02288* remained nominally significant in the AFR tract and standard GWAS (p<0.001). Interestingly, SNPs in *LRP1B* were also identified in the EUR tract. Because the LD between rs1968911 and EUR tract lead SNP rs78585608 is nearly zero in these participants (r^2^=0.0029), these likely represent two distinct signals, one in the AFR component and one in the EUR component. The previous association of rs7526762 in FPGT-TNNI3K/TNNI3K failed to replicate (p>0.05). Importantly, the *APOL1* G1 variants implicated in various kidney diseases^84–87^ including SCDN^22,88–90^ were nominally associated with eGFR in the AFR tract (p<0.01) but not the EUR tract (p=0.9). This is unsurprising, given how rare the G1 variants are in Europeans. Much like the array-based GWAS, this finding confirms that variation in *APOL1*, while associated, is not the strongest genetic risk factor for SCDN.

There are several important differences between the current analysis and the previous array-based GWAS study. First, we utilized WGS instead of array data, thus providing the opportunity for novel discoveries. Second, we estimated GFR with the newer CKD-EPI-2021 creatinine-based formula (without adjustment for race) as opposed to the previously used Modification of Diet in Renal Disease (MDRD) equation, which requires more laboratory measures and therefore reduced our sample size. The correlation between eGFR based on MDRD and CKD-EPI-2021 in OMG-SCD was 89%, demonstrating strong concordance between the two estimates. However, both equations are known to overestimate GFR in SCD patients due to increased glomerular filtration, lower muscle mass, and increased dysfunction of tubular secretion, therefore underestimating the prevalence of CKD^91–93^. Future studies should compare the performance of creatinine-based equations with that of cystatin-C based equations or measured GFR.

While this study represents the largest WGS analysis of SCDN to date, we remain underpowered to detect small effects. Therefore, we chose to examine only biallelic SNPs with frequency above 1%. Efforts to collect larger SCD cohorts should be prioritized to replicate these findings and investigate potential targets for interventions. Future work should also interrogate the contribution of more rare variants and of structural variants to SCDN.

In summary, this first local ancestry-aware GWAS of kidney function in SCD patients has identified many GWS regions encompassing genes and pathways previously implicated in renal dysfunction or SCD pathology. While most eGFR-associated regions in the AFR component mirrored previous standard GWAS, we were nonetheless able to identify one unique locus in the AFR tract that was missed in the standard GWAS. Perhaps more importantly, we were able to examine the EUR contribution to risk, which is largely masked in traditional approaches due to a low global percentage of admixture. Interestingly, many of the eGFR-associated genes and pathways identified are also integrally related to SCD pathophysiology. This analysis furthers our understanding of the genetic underpinnings of SCDN and provides additional targets for therapeutics aimed at curbing the impact of kidney disease in SCD.

## Data Availability

The datasets used in this analysis are available via controlled access in the database of Genotypes and Phenotypes (dbGaP) under accession number phs001608 for OMG-SCD and phs001514 for Walk-PHaSST. For questions regarding the OMG-SCD cohort, please contact; for Walk-PHaSST contact.

## ACKNOWLEDGEMENTS

The authors thank all the subjects for participating in this study. This work was supported in part by grant awards 2015131 and 2012126 from the Doris Duke Charitable Foundation, R01HL68959 and R01HL079915 from the National Heart, Lung and Blood Institute, R21AG0849 from the National Institute on Aging, and R01DK110104 and R21DK124836 from the National Institute of Diabetes and Digestive and Kidney Diseases. The Walk-PHaSST cohort was collected as a clinical trial (NCT00492531) funded with federal funds from the NHLBI, NIH, and Department of Health and Human Services under contract HHSN268200617182C. Molecular data for the TOPMed program was supported by NHLBI; whole genome sequencing for OMG-SCD (phs001608) and Walk-PHaSST (phs001514) was performed at Baylor College of Medicine Human Genome Sequencing Center (HHSN268201500015C). Core support including centralized genomic read mapping and genotype calling, along with variant quality metrics and filtering were provided by the TOPMed Informatics Research Centre (3R01HL-117626-02S1; contract HHSN268201800002I). Core support including phenotype harmonization, data management, sample-identity QC, and general program coordination was provided by the TOPMed Data Coordinating Centre (R01HL-120393; U01HL-120393; contract HHSN268201800001I).

## AUTHORSHIP CONTRIBUTIONS

AAK and MJT designed the research study. VRG and MTG generated the Walk-PHaSST data. MEG, AAK, and MJT analyzed the data. MEG drafted the manuscript. All authors provided edits to and approved the final version of the manuscript.

## REFERENCES

1. Hassell KL. Population estimates of sickle cell disease in the U.S. Am J Prev Med. 2010;38(4 Suppl):S512–521.

2. Telen MJ, Malik P, Vercellotti GM. Therapeutic strategies for sickle cell disease: towards a multi-agent approach. Nat Rev Drug Discov. 2019;18(2):139–158.

3. Elmariah H, Garrett ME, De Castro LM, et al. Factors associated with survival in a contemporary adult sickle cell disease cohort. Am J Hematol. 2014;89(5):530–535.

4. Fitzhugh CD, Lauder N, Jonassaint JC, et al. Cardiopulmonary complications leading to premature deaths in adult patients with sickle cell disease. Am J Hematol. 2010;85(1):36–40.

5. Platt OS, Brambilla DJ, Rosse WF, et al. Mortality in sickle cell disease. Life expectancy and risk factors for early death. N Engl J Med. 1994;330(23):1639–1644.

6. Powars DR, Chan LS, Hiti A, Ramicone E, Johnson C. Outcome of sickle cell anemia: a 4-decade observational study of 1056 patients. Medicine (Baltimore). 2005;84(6):363–376.

7. Hamideh D, Alvarez O. Sickle cell disease related mortality in the United States (1999-2009). Pediatr Blood Cancer. 2013;60(9):1482–1486.

8. McClellan AC, Luthi JC, Lynch JR, et al. High one year mortality in adults with sickle cell disease and end-stage renal disease. Br J Haematol. 2012;159(3):360–367.

9. Guasch A, Navarrete J, Nass K, Zayas CF. Glomerular involvement in adults with sickle cell hemoglobinopathies: Prevalence and clinical correlates of progressive renal failure. J Am Soc Nephrol. 2006;17(8):2228–2235.

10. Powars DR, Elliott-Mills DD, Chan L, et al. Chronic renal failure in sickle cell disease: risk factors, clinical course, and mortality. Ann Intern Med. 1991;115(8):614–620.

11. Morris AP, Le TH, Wu H, et al. Trans-ethnic kidney function association study reveals putative causal genes and effects on kidney-specific disease aetiologies. Nat Commun. 2019;10(1):29.

12. Teumer A, Li Y, Ghasemi S, et al. Genome-wide association meta-analyses and fine-mapping elucidate pathways influencing albuminuria. Nat Commun. 2019;10(1):4130.

13. Wuttke M, Li Y, Li M, et al. A catalog of genetic loci associated with kidney function from analyses of a million individuals. Nat Genet. 2019;51(6):957–972.

14. Stanzick KJ, Li Y, Schlosser P, et al. Discovery and prioritization of variants and genes for kidney function in >1.2 million individuals. Nat Commun. 2021;12(1):4350.

15. Garrett ME, Soldano KL, Erwin KN, et al. Genome-wide meta-analysis identifies new candidate genes for sickle cell disease nephropathy. Blood Adv. 2023;7(17):4782–4793.

16. Kamiza AB, Chikowore T, Chen G, et al. KidneyGenAfrica multi-cohort Genome-wide association study and polygenic prediction of kidney function in 110,000 Africans. Nat Commun. 2026.

17. Reynolds KM, Pasteris J, Best LG, et al. Genome-wide association of albuminuria and chronic kidney disease in American Indians and Hispanics/Latinos. Hum Mol Genet. 2026;35(1).

18. Fatumo S, Chikowore T, Kalyesubula R, et al. Discovery and fine-mapping of kidney function loci in first genome-wide association study in Africans. Hum Mol Genet. 2021;30(16):1559–1568.

19. Collaborators GBDSCD. Global, regional, and national prevalence and mortality burden of sickle cell disease, 2000-2021: a systematic analysis from the Global Burden of Disease Study 2021. Lancet Haematol. 2023;10(8):e585–e599.

20. Shriner D. Overview of admixture mapping. Curr Protoc Hum Genet. 2013;Chapter 1:Unit 1 23.

21. Atkinson EG, Maihofer AX, Kanai M, et al. Tractor uses local ancestry to enable the inclusion of admixed individuals in GWAS and to boost power. Nat Genet. 2021;53(2):195–204.

22. Ashley-Koch AE, Okocha EC, Garrett ME, et al. MYH9 and APOL1 are both associated with sickle cell disease nephropathy. Br J Haematol. 2011;155(3):386–394.

23. Machado RF, Barst RJ, Yovetich NA, et al. Hospitalization for pain in patients with sickle cell disease treated with sildenafil for elevated TRV and low exercise capacity. Blood. 2011;118(4):855–864.

24. Inker LA, Eneanya ND, Coresh J, et al. New Creatinine- and Cystatin C-Based Equations to Estimate GFR without Race. N Engl J Med. 2021;385(19):1737–1749.

25. Taliun D, Harris DN, Kessler MD, et al. Sequencing of 53,831 diverse genomes from the NHLBI TOPMed Program. Nature. 2021;590(7845):290–299.

26. Purcell S, Neale B, Todd-Brown K, et al. PLINK: a tool set for whole-genome association and population-based linkage analyses. Am J Hum Genet. 2007;81(3):559–575.

27. Price AL, Patterson NJ, Plenge RM, Weinblatt ME, Shadick NA, Reich D. Principal components analysis corrects for stratification in genome-wide association studies. Nat Genet. 2006;38(8):904–909.

28. Falush D, Stephens M, Pritchard JK. Inference of population structure using multilocus genotype data: linked loci and correlated allele frequencies. Genetics. 2003;164(4):1567–1587.

29. Tandon A, Patterson N, Reich D. Ancestry informative marker panels for African Americans based on subsets of commercially available SNP arrays. Genet Epidemiol. 2011;35(1):80–83.

30. Maples BK, Gravel S, Kenny EE, Bustamante CD. RFMix: a discriminative modeling approach for rapid and robust local-ancestry inference. Am J Hum Genet. 2013;93(2):278–288.

31. Willer CJ, Li Y, Abecasis GR. METAL: fast and efficient meta-analysis of genomewide association scans. Bioinformatics. 2010;26(17):2190–2191.

32. Viechtbauer W. Conducting Meta-Analyses in R with the metafor Package. Journal of Statistical Software. 2010;36(3):1–48.

33. Turner SD. qqman: an R package for visualizing GWAS results using Q-Q and manhattan plots. Journal of Open Source Software. 2018;3(25):731.

34. Wang K, Li M, Hakonarson H. ANNOVAR: functional annotation of genetic variants from high-throughput sequencing data. Nucleic Acids Res. 2010;38(16):e164.

35. Hinrichs AS, Karolchik D, Baertsch R, et al. The UCSC Genome Browser Database: update 2006. Nucleic Acids Res. 2006;34(Database issue):D590–598.

36. Watanabe K, Taskesen E, van Bochoven A, Posthuma D. Functional mapping and annotation of genetic associations with FUMA. Nat Commun. 2017;8(1):1826.

37. Kircher M, Witten DM, Jain P, O’Roak BJ, Cooper GM, Shendure J. A general framework for estimating the relative pathogenicity of human genetic variants. Nat Genet. 2014;46(3):310–315.

38. Han SK, McNulty MT, Benway CJ, et al. Mapping genomic regulation of kidney disease and traits through high-resolution and interpretable eQTLs. Nat Commun. 2023;14(1):2229.

39. Martin V, Zhao J, Afek A, Mielko Z, Gordan R. QBiC-Pred: quantitative predictions of transcription factor binding changes due to sequence variants. Nucleic Acids Res. 2019;47(W1):W127–W135.

40. de Leeuw CA, Mooij JM, Heskes T, Posthuma D. MAGMA: generalized gene-set analysis of GWAS data. PLoS Comput Biol. 2015;11(4):e1004219.

41. Storey JD BA, Dabney A, Robinson D. qvalue: Q-value estimation for false discovery rate control. R package version 2420. 2025.

42. Ge SX, Jung D, Yao R. ShinyGO: a graphical gene-set enrichment tool for animals and plants. Bioinformatics. 2020;36(8):2628–2629.

43. Davis TL, Walker JR, Campagna-Slater V, et al. Structural and biochemical characterization of the human cyclophilin family of peptidyl-prolyl isomerases. PLoS Biol. 2010;8(7):e1000439.

44. Hadpech S, Thongboonkerd V. Current update on theranostic roles of cyclophilin A in kidney diseases. Theranostics. 2022;12(9):4067–4080.

45. Wu S, Zou Y, Tan X, et al. The molecular mechanisms of peptidyl-prolyl cis/trans isomerase Pin1 and its relevance to kidney disease. Front Pharmacol. 2024;15:1373446.

46. Rane MJ, Zhao Y, Cai L. Krupsilonppel-like factors (KLFs) in renal physiology and disease. EBioMedicine. 2019;40:743–750.

47. Hayashi K, Sasamura H, Nakamura M, et al. KLF4-dependent epigenetic remodeling modulates podocyte phenotypes and attenuates proteinuria. J Clin Invest. 2014;124(6):2523–2537.

48. Francis A BJ, Francis L, McTaggart S, Mallett A. Polyploid Change of the Glomerular Basement Membrane in a Child with Steroid Resistant Nephrotic Syndrome and ARHGAP24 Mutation: A Case Report. The Open Urology & Nephrology Journal. 2016.

49. Akilesh S, Suleiman H, Yu H, et al. Arhgap24 inactivates Rac1 in mouse podocytes, and a mutant form is associated with familial focal segmental glomerulosclerosis. J Clin Invest. 2011;121(10):4127–4137.

50. Ayari F, Abdollahzade Fard A, Chodari L. Selenium pretreatment improve renal function, autophagy signaling pathway and mir21a gene expression in renal ischemia reperfusion injury model in male rat. J Trace Elem Med Biol. 2025;88:127610.

51. Liu X, Hong Q, Wang Z, Yu Y, Zou X, Xu L. MiR-21 inhibits autophagy by targeting Rab11a in renal ischemia/reperfusion. Exp Cell Res. 2015;338(1):64–69.

52. Ye CL, Du Y, Yu X, et al. STEAP3 Affects Ferroptosis and Progression of Renal Cell Carcinoma Through the p53/xCT Pathway. Technol Cancer Res Treat. 2022;21:15330338221078728.

53. Wu J, Bi Q, Zheng X, et al. STEAP3 can predict the prognosis and shape the tumor microenvironment of clear cell renal cell carcinoma. BMC Cancer. 2022;22(1):1204.

54. Zhang F, Tao Y, Zhang Z, et al. Metalloreductase Steap3 coordinates the regulation of iron homeostasis and inflammatory responses. Haematologica. 2012;97(12):1826–1835.

55. Li S, Han Q, Liu C, et al. Role of ferroptosis in chronic kidney disease. Cell Commun Signal. 2024;22(1):113.

56. Guo X, Engel JL, Xiao J, et al. UBLCP1 is a 26S proteasome phosphatase that regulates nuclear proteasome activity. Proc Natl Acad Sci U S A. 2011;108(46):18649–18654.

57. Makino SI, Shirata N, Oliva Trejo JA, et al. Impairment of Proteasome Function in Podocytes Leads to CKD. J Am Soc Nephrol. 2021;32(3):597–613.

58. Shin YJ, Jeon YJ, Jung N, Park JW, Park HY, Jung SC. Substrate-specific gene expression profiles in different kidney cell types are associated with Fabry disease. Mol Med Rep. 2015;12(4):5049–5057.

59. Wu G, Li J, Xu Y, Che X, Chen F, Wang Q. A New Survival Model Based on ADAMTSs for Prognostic Prediction in Clear Cell Renal Cell Carcinoma. J Oncol. 2021;2021:2606213.

60. Schlosser P, Tin A, Matias-Garcia PR, et al. Meta-analyses identify DNA methylation associated with kidney function and damage. Nat Commun. 2021;12(1):7174.

61. Smyth LJ, Kerr KR, Kilner J, McGill AE, Maxwell AP, McKnight AJ. Longitudinal Epigenome-Wide Analysis of Kidney Transplant Recipients Pretransplant and Posttransplant. Kidney Int Rep. 2023;8(2):330–340.

62. Weck ML, Crawley SW, Stone CR, Tyska MJ. Myosin-7b Promotes Distal Tip Localization of the Intermicrovillar Adhesion Complex. Curr Biol. 2016;26(20):2717–2728.

63. Ashworth SL, Molitoris BA. Pathophysiology and functional significance of apical membrane disruption during ischemia. Curr Opin Nephrol Hypertens. 1999;8(4):449–458.

64. Elmubarak I, Shril S, Mansour B, et al. Recessive variants in MYO1C as a potential novel cause of proteinuric kidney disease. Pediatr Nephrol. 2024;39(10):2939–2945.

65. Ostendorf T, Boor P, van Roeyen CR, Floege J. Platelet-derived growth factors (PDGFs) in glomerular and tubulointerstitial fibrosis. Kidney Int Suppl (2011). 2014;4(1):65–69.

66. Okamoto K, Tokunaga K, Doi K, et al. Common variation in GPC5 is associated with acquired nephrotic syndrome. Nat Genet. 2011;43(5):459–463.

67. Okamoto K, Honda K, Doi K, et al. Glypican-5 Increases Susceptibility to Nephrotic Damage in Diabetic Kidney. Am J Pathol. 2015;185(7):1889–1898.

68. Gee HY, Zhang F, Ashraf S, et al. KANK deficiency leads to podocyte dysfunction and nephrotic syndrome. J Clin Invest. 2015;125(6):2375–2384.

69. Oda K, Katayama K, Zang L, et al. The Protective Role of KANK1 in Podocyte Injury. Int J Mol Sci. 2024;25(11).

70. Zhang L, Xu P, Hao L, Wang L, Xu Y, Jiang C. The role of transient receptor potential channels in chronic kidney disease-mineral and bone disorder. Front Pharmacol. 2025;16:1583487.

71. Rodelo-Haad C, Pendon-Ruiz de Mier MV, Diaz-Tocados JM, et al. The Role of Disturbed Mg Homeostasis in Chronic Kidney Disease Comorbidities. Front Cell Dev Biol. 2020;8:543099.

72. Peroumal D, Jawale CV, Choi W, et al. The survival of B cells is compromised in kidney disease. Nat Commun. 2024;15(1):10842.

73. Taguchi S, Azushima K, Yamaji T, et al. Effects of tumor necrosis factor-alpha inhibition on kidney fibrosis and inflammation in a mouse model of aristolochic acid nephropathy. Sci Rep. 2021;11(1):23587.

74. Ma HY, Chen S, Du Y. Estrogen and estrogen receptors in kidney diseases. Ren Fail. 2021;43(1):619–642.

75. Ballas SK, Mohandas N. Sickle red cell microrheology and sickle blood rheology. Microcirculation. 2004;11(2):209–225.

76. George A, Pushkaran S, Li L, et al. Altered phosphorylation of cytoskeleton proteins in sickle red blood cells: the role of protein kinase C, Rac GTPases, and reactive oxygen species. Blood Cells Mol Dis. 2010;45(1):41–45.

77. George A, Pushkaran S, Konstantinidis DG, et al. Erythrocyte NADPH oxidase activity modulated by Rac GTPases, PKC, and plasma cytokines contributes to oxidative stress in sickle cell disease. Blood. 2013;121(11):2099–2107.

78. Cain SA, Woods S, Singh M, Kimber SJ, Baldock C. ADAMTS6 cleaves the large latent TGFbeta complex and increases the mechanotension of cells to activate TGFbeta. Matrix Biol. 2022;114:18–34.

79. Franco-Penteado CF HS, Conran N, Saad STO, Costa FF. Increased Levels and Activities of Matrix Metalloproteinases in Sickle Cell Disease. Blood. 2006;108(11):1220.

80. Torres Lde S, Okumura JV, da Silva DG, et al. Plasma levels of TGF-beta1 in homeostasis of the inflammation in sickle cell disease. Cytokine. 2016;80:18–25.

81. Leslie TM P-ST. Hemolysis Induced Induction of Matrix Metalloproteinases in Sickle Cell Disease. Blood. 2023;142:5284.

82. Liu L, Pertsemlidis A, Ding LH, et al. Original Research: A case-control genome-wide association study identifies genetic modifiers of fetal hemoglobin in sickle cell disease. Exp Biol Med (Maywood). 2016;241(7):706–718.

83. Wongborisuth C, Chumchuen S, Sripichai O, et al. Down-regulation of the transcriptional repressor ZNF802 (JAZF1) reactivates fetal hemoglobin in beta(0)-thalassemia/HbE. Sci Rep. 2022;12(1):4952.

84. Freedman BI, Kopp JB, Langefeld CD, et al. The apolipoprotein L1 (APOL1) gene and nondiabetic nephropathy in African Americans. J Am Soc Nephrol. 2010;21(9):1422–1426.

85. Genovese G, Friedman DJ, Ross MD, et al. Association of trypanolytic ApoL1 variants with kidney disease in African Americans. Science. 2010;329(5993):841–845.

86. Kopp JB, Nelson GW, Sampath K, et al. APOL1 genetic variants in focal segmental glomerulosclerosis and HIV-associated nephropathy. J Am Soc Nephrol. 2011;22(11):2129–2137.

87. Tzur S, Rosset S, Shemer R, et al. Missense mutations in the APOL1 gene are highly associated with end stage kidney disease risk previously attributed to the MYH9 gene. Hum Genet. 2010;128(3):345–350.

88. Saraf SL, Zhang X, Shah B, et al. Genetic variants and cell-free hemoglobin processing in sickle cell nephropathy. Haematologica. 2015;100(10):1275–1284.

89. Schaefer BA, Flanagan JM, Alvarez OA, et al. Genetic Modifiers of White Blood Cell Count, Albuminuria and Glomerular Filtration Rate in Children with Sickle Cell Anemia. PLoS One. 2016;11(10):e0164364.

90. Geard A, Pule GD, Chetcha Chemegni B, et al. Clinical and genetic predictors of renal dysfunctions in sickle cell anaemia in Cameroon. Br J Haematol. 2017;178(4):629–639.

91. Asnani MR, Lynch O, Reid ME. Determining glomerular filtration rate in homozygous sickle cell disease: utility of serum creatinine based estimating equations. PLoS One. 2013;8(7):e69922.

92. Afangbedji N, Jerebtsova M. Glomerular filtration rate abnormalities in sickle cell disease. Front Med (Lausanne). 2022;9:1029224.

93. Derebail VK ZL, Elsherif L, Patillo KL, Wichlan D, Landes K, McCune P, Loehr L, Cronin RM, Desai PC, Cai J, Ataga KI. Evaluating Equations for Estimated Glomerular Filtration Rate (eGFR) in Patients with Sickle Cell Disease (SCD). Blood. 2022;140:2545–2546.

