## Supplemental Figures and Table legends for "Local ancestry-aware genome-wide meta-analysis uncovers novel genetic loci for sickle cell disease nephropathy"

**Table S1**. Cohort-specific results for African tract genome-wide significant SNPs

**Table S2**. Cohort-specific results for European tract genome-wide significant SNPs

**Table S3**. African tract genome-wide significant SNP results including evidence of regulatory impact (CADD scores, RegulomeDB scores and transcription factor binding disruption)

**Table S4**. European tract genome-wide significant SNP results including evidence of regulatory impact (CADD scores, RegulomeDB scores and transcription factor binding disruption)

**Table S5**. Genes mapped to African tract genome-wide significant SNPs with probability of loss-of-function intolerance p-value (pLI)

**Table S6**. Genes mapped to European tract genome-wide significant SNPs with probability of loss-of-function intolerance p-value (pLI)

**Table S7**. Standard GWAS genome-wide significant regions

**Table S8**. Standard GWAS genome-wide significant SNP results including evidence of regulatory impact (CADD scores, RegulomeDB scores and transcription factor binding disruption)

**Table S9**. Genes mapped to standard GWAS genome-wide significant SNPs with probability of loss-of-function intolerance p-value (pLI)

**Table S10**. MAGMA gene-based results for the European tract ($q<0.05$)

**Table S11**. Gene set enrichment results ($q<0.05$) for the MAGMA gene-based analysis ($p<0.01$) in the European tract

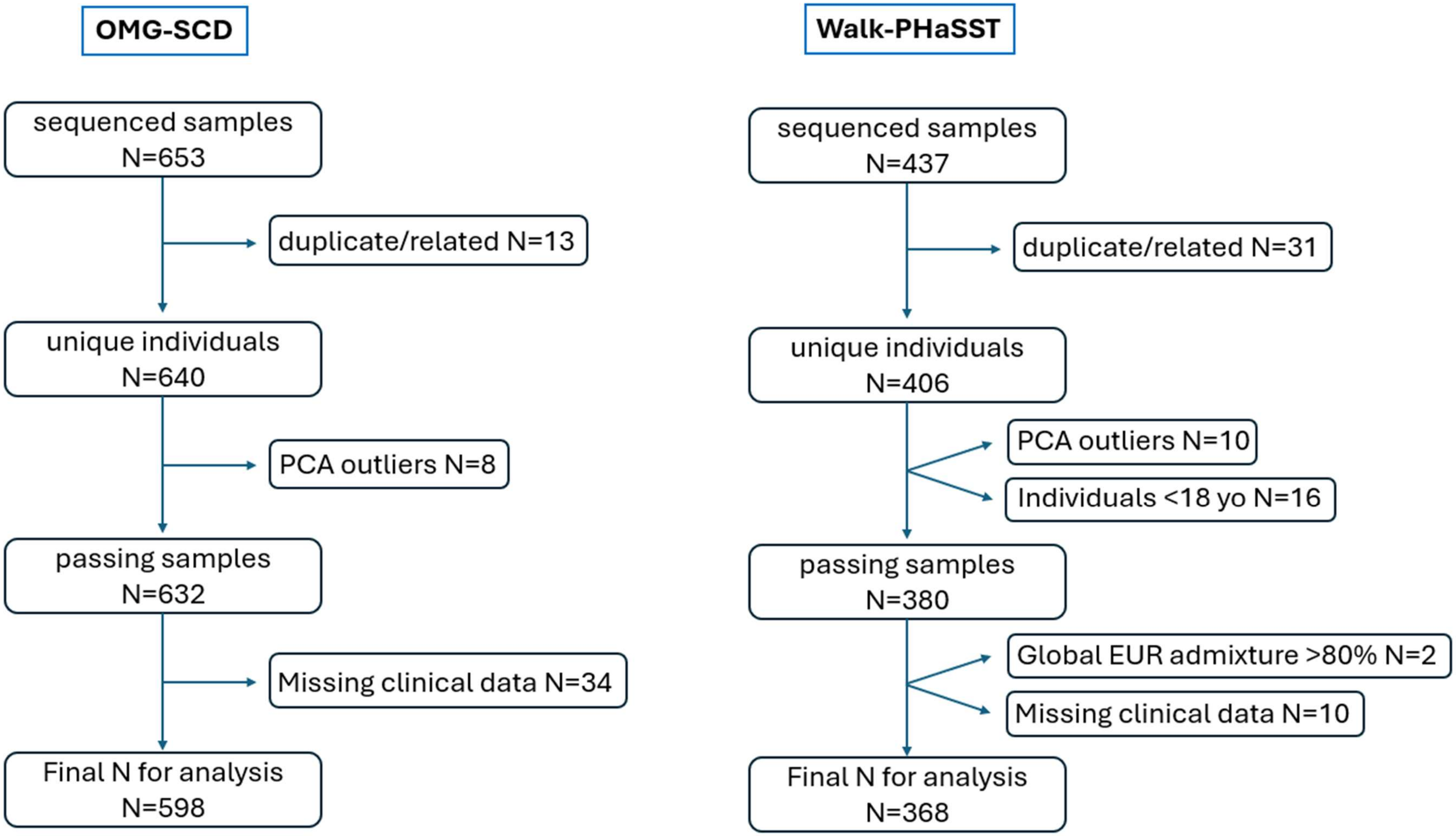


**Figure S1**. CONSORT style diagram depicting the sample exclusions made in OMG-SCD and Walk-PHaSST from sequenced samples to those included in statistical analysis.

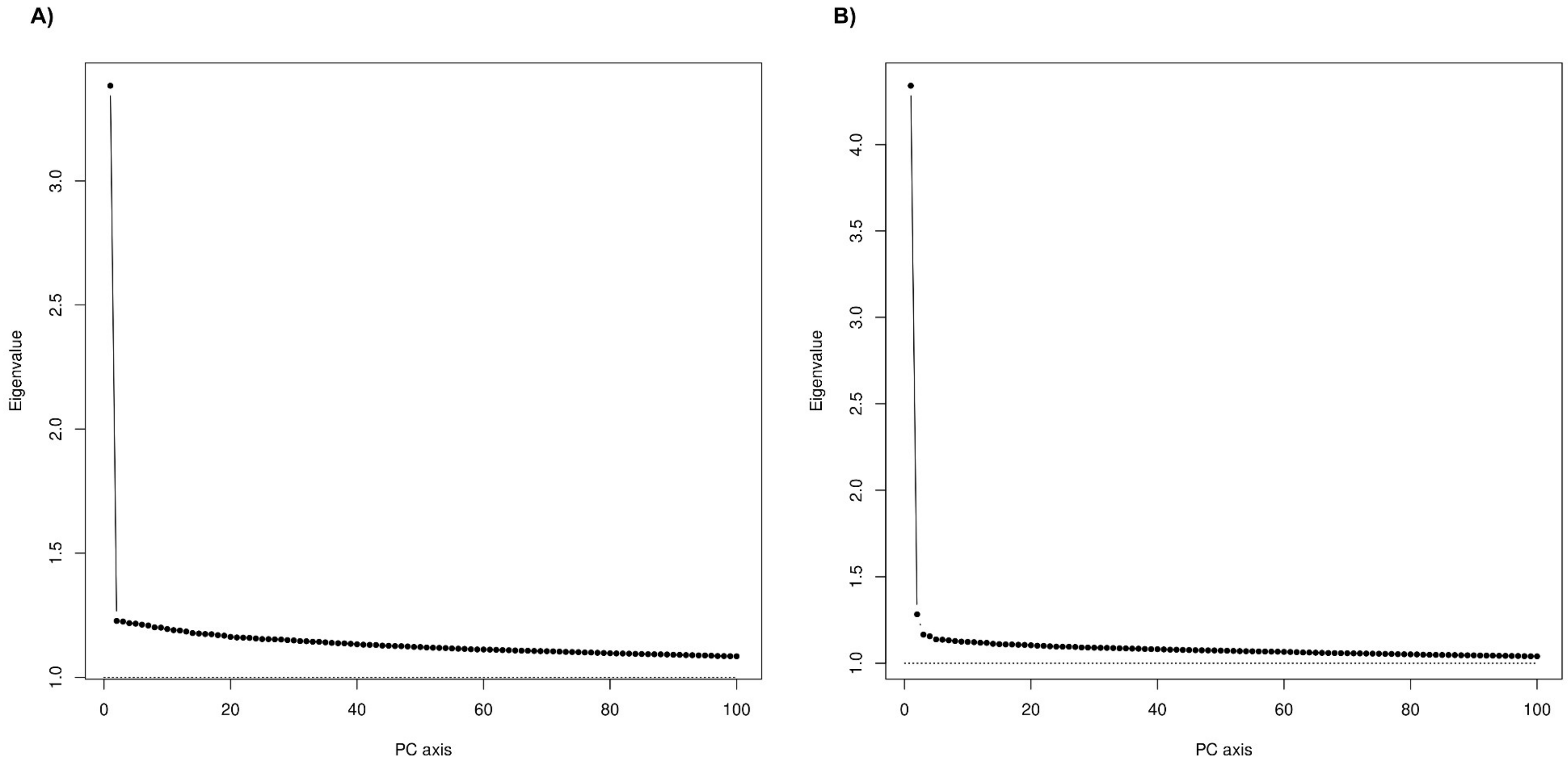


**Figure S2**. Scree plots of eigenvalues from principal component (PC) analysis of A) OMG-SCD participants and B) Walk-PHaSST participants. Visual inspection indicates that two PCs and five PCs are necessary to control for population substructure in OMG-SCD and Walk-PHaSST, respectively.

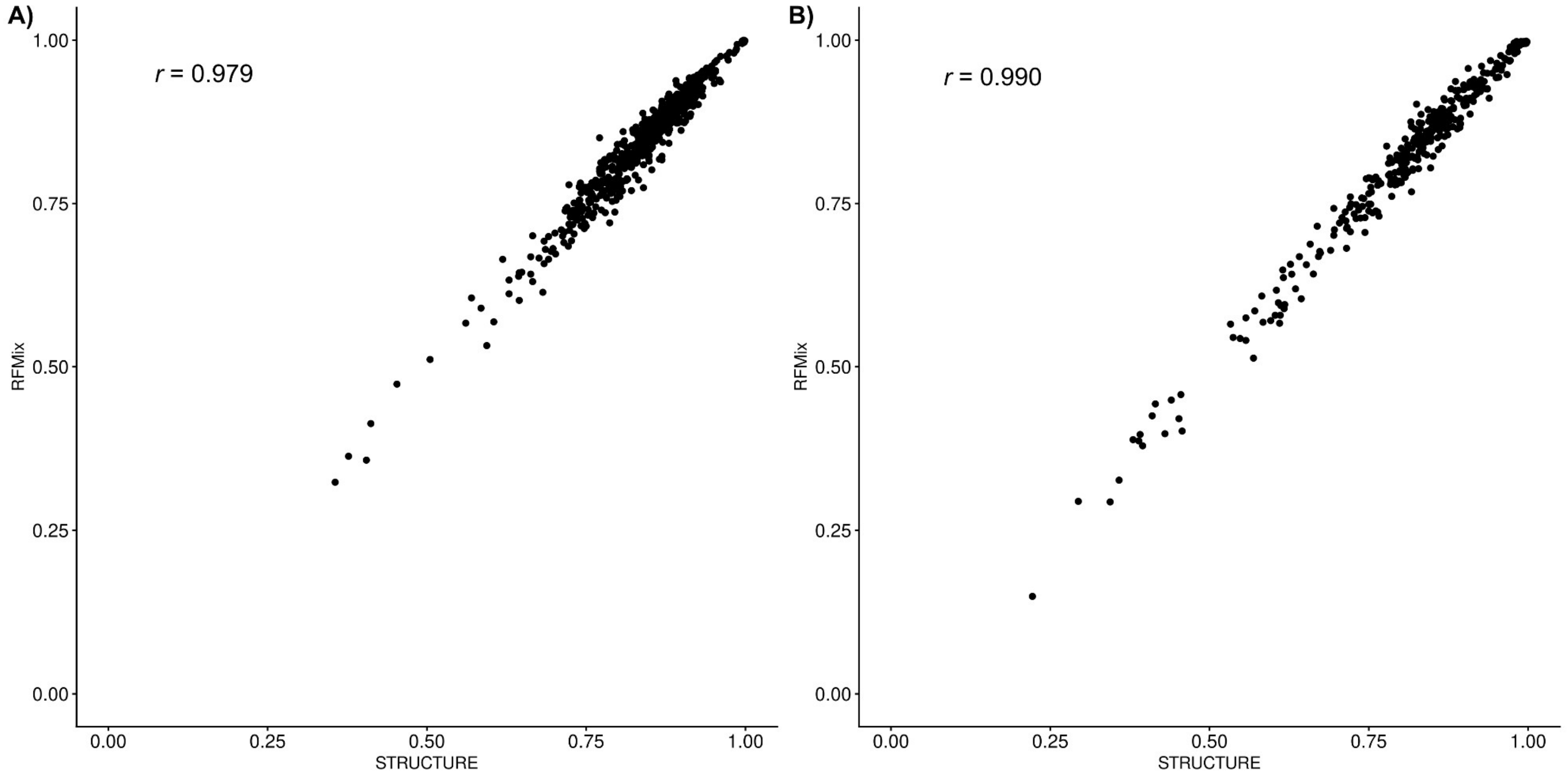


**Figure S3**. Correlation of AFR global admixture proportion estimates obtained from STRUCTURE compared to genome-wide averages of local ancestry estimates from RFMix for A) OMG-SCD and B) Walk-PHaSST.

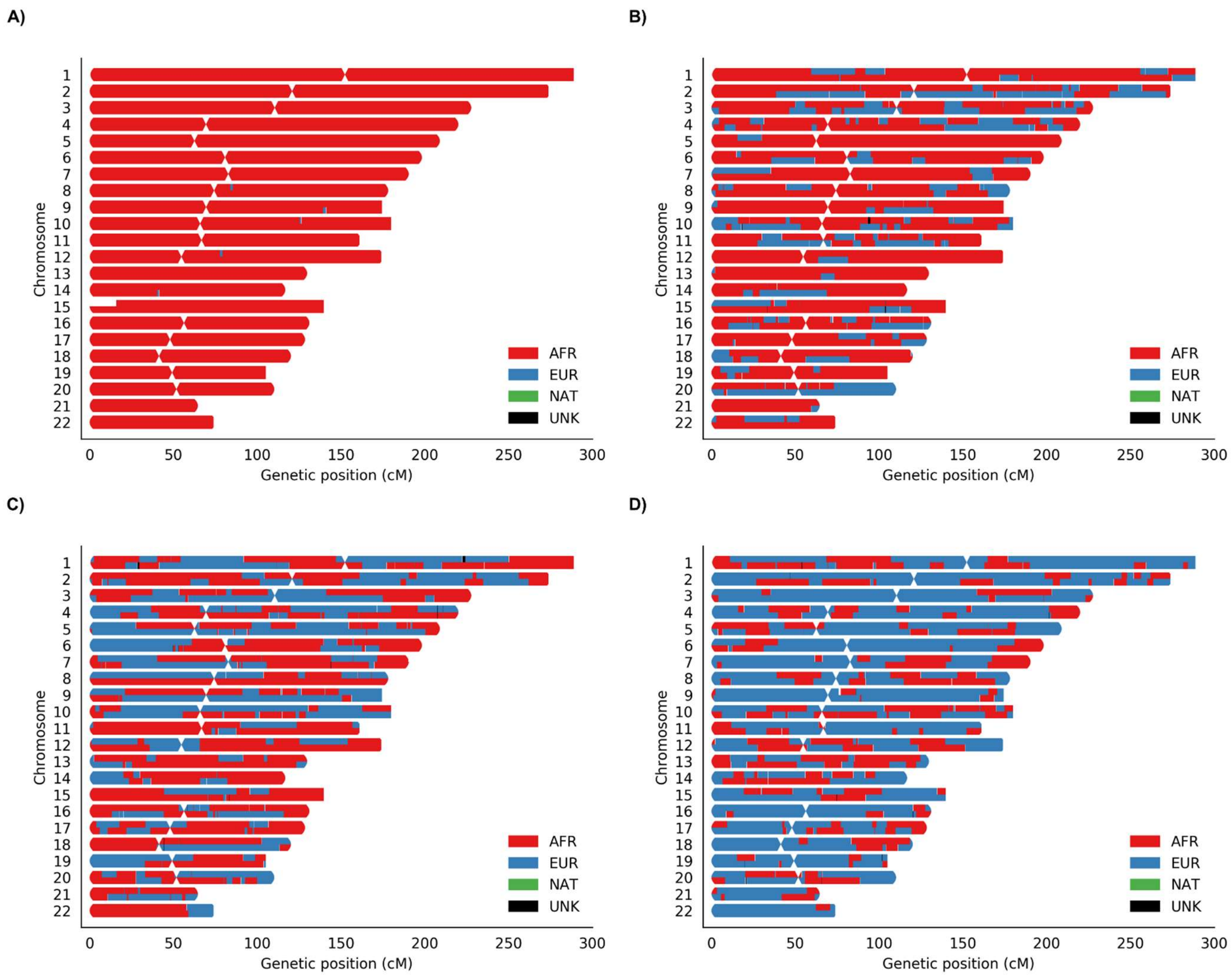


**Figure S4**. Painted karyogram plots for four selected OMG-SCD participants with varying proportions of global European (EUR) admixture including A) EUR admixture=0.002 B) EUR admixture=0.254 C) EUR admixture=0.415 and D) EUR admixture=0.677.

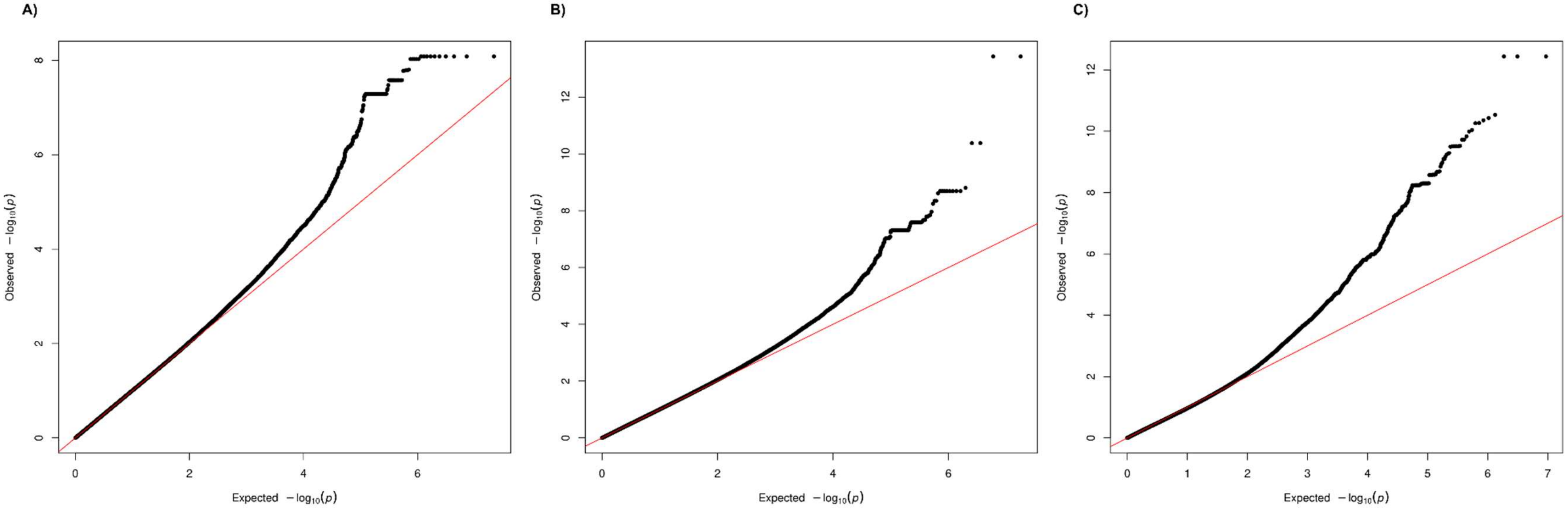


**Figure S5**. Quantile-quantile (QQ) plots for the A) standard eGFR GWAS, B) the AFR tract eGFR GWAS, and (C) the EUR tract eGFR GWAS. The genomic inflation factor (λ) was 1.007, 1.015, and 0.967, respectively.
